# Adherence to the EAT-Lancet diet and risk of stroke and stroke subtypes: A Danish cohort study

**DOI:** 10.1101/2021.04.14.21255371

**Authors:** Daniel B. Ibsen, Anne H. Christiansen, Anja Olsen, Anne Tjønneland, Kim Overvad, Alicja Wolk, Janne K. Mortensen, Christina C. Dahm

## Abstract

**Objective:** To investigate the association between adherence to the EAT-Lancet diet, a sustainable and mostly plant-based diet, and risk of stroke and subtypes of stroke in a Danish population. For comparison, we also investigated the Alternate Healthy Eating Index-2010 (AHEI).

**Methods:** We used the Danish Diet, Cancer and Health cohort (n=55,016) including adults aged 50-64 years at baseline (1993-1997). A food frequency questionnaire was used to assess dietary intake and group participants according to adherence to the diets. Stroke cases were identified using a national registry and subsequently validated by review of medical records (n=2253). Cox proportional hazards models were used to estimate hazard ratios (HR) and 95% confidence intervals (CI) for associations with the EAT-Lancet diet or the AHEI and risk of stroke and stroke subtypes.

**Results:** Adherence to the EAT-Lancet diet was associated with a lower risk of stroke, although not statistically significant (highest vs lowest adherence: HR 0.91; 95% CI 0.76, 1.09). A lower risk was observed for the AHEI (0.75; 0.64, 0.87). For stroke subtypes we found that adherence to the EAT-Lancet diet was associated with a lower risk of subarachnoid hemorrhage (0.30; 0.12, 0.73) and the AHEI was associated with a lower risk of ischemic stroke (0.76; 0.64, 0.90) and intracerebral hemorrhage (0.58; 0.36, 0.93).

**Conclusions:** Adherence to the EAT-Lancet diet was associated with a lower risk of subarachnoid stroke and the AHEI was associated with a lower risk of total stroke, mainly ischemic stroke and intracerebral hemorrhage.

## Introduction

Our diets are negatively affecting both planetary and population health.^1^ Shifting population dietary habits towards more sustainable foods is critical to reduce the detrimental effects of human activity on the planet, but this shift must not have the unintended side effect of harming population health. The EAT-Lancet Commission on Food, Planet and Health called for a global transformation of the food system to address human and planetary health, and proposed a global reference diet, including mainly plant-based foods, limited dairy products and little meat, that at a planetary level represents the best evidence to date for a sustainable healthy diet.^2^

Previous results from the European Prospective Investigation into Cancer and nutrition (EPIC)-Oxford study suggested that vegetarians, consuming plant-based foods, some dairy products, and no meat, had a higher risk of total and hemorrhagic stroke compared with meat eaters.^3^ This finding indicates that there could be potential unintended side effects of following a plant-based diet. In contrast, results from three US cohorts of health professionals suggested that adherence to the healthy plant-based diet index was associated with a lower risk of total stroke, but with uncertainty in analyses of stroke subtypes, likely due to too few cases.^4^ Several nutrients in the diets consumed may play a role in stroke etiology; some may lower the risk like dietary fibre, omega-3 fatty acids, and potassium while others may elevelevate the risk, such as cholesterol, saturated fat, calcium, sodium, and heme iron.^5^ As dietary patterns contain different amounts of these nutrients, they likely exert varying associations with stroke risk. Stroke is a heterogenous disease with different etiologies. Few studies have, however, related different dietary patterns to development of stroke subtypes, particularly different types of hemorrhagic stroke. Taken together, this leaves some uncertainty about the effect of transitioning to a plant-based diet and risk of stroke and its subtypes.

We aimed to investigate the association between adherence to the EAT-Lancet diet and risk of total and subtypes of stroke in a Danish population. We also investigated the association between adherence to the Alternate Healthy Eating Index-2010 (AHEI) and risk of stroke as this diet score broadly reflects dietary guidelines without a focus on sustainability. As some population subgroups may experience greater benefits from adopting a plant-based diet than others, in terms of affecting the risk of stroke, we also investigated effect modification by age, sex and body mass index (BMI), on the additive scale.

## Methods

### Study population

We used data from the Danish Diet, Cancer and Health cohort. In December 1993 to May 1997, all adults aged 50-64 years, residing the greater Copenhagen and Aarhus areas and without any previous diagnosis of cancer registered in the Danish Cancer Registry were invited to participate in the study (n=160,725). In total, 57,053 participated in the study. Participants completed a food frequency questionnaire (FFQ) before visiting one of two study centers, where they completed a lifestyle questionnaire and had a standardized physical examination conducted by trained personnel. All questionnaires were checked by the personnel at the study centers.

For this study we excluded those with a previous diagnosis of cancer (due to a delay in registries when sending the invitations to participate) (n=585), a previous diagnosis of stroke (n=582) or missing information in covariates (n=870). In total 55,016 participants were included in the analyses (eFigure 1).

### Standard protocol approvals, registrations and patient consents

The Danish Diet, Cancer and Health cohort was approved by the National Committee on Health Research Ethics (jr.nr. (KF) 01-345-93). This project was additionally approved by the IRB at the Danish Cancer Society and registered internally at Aarhus University in order to comply with the GDPR and Danish Data Protection Act. All participants gave written informed consent to participate.

### Diet assessment

The participants completed a 192-item semiquantitative FFQ designed for the study^6^ to reflect the average intake in the previous 12 months. Response options ranged from never to 8 times or more per day, and responses were converted into daily intake of foods and nutrients using the software FoodCalc^7^ and the Danish National Food Tables.^8^ The FFQ was validated against two 7-day food records.^9^ The agreement between the FFQ and food records for protein, carbohydrates and fat for classification of participants into quintiles of intake was 65%, 74% and 75% respectively.^9^ Because the FFQ was designed for a Danish population, it posed detailed questions about intake of grains, potatoes, vegetables, fruits, dairy products, meat products and snacks. The intake of legumes was very low in the population and hence only intake of peanuts (as a snack) was included as a specific question.

We created the EAT-Lancet diet score similarly to Knuppel et al.^10^, based on the guidelines set by the EAT-Lancet Commission.^2^ The score includes 14 dietary components from 9 overall food categories: Rice, grains and corn including whole grains, tubers and starchy vegetables, vegetables, fruits, dairy products, protein sources, legumes, added fats and added sugar. Adherence to each component was given 1 point, and non-adherence 0 points. The EAT-Lancet diet score is the sum of points for each individual. Further details are provided in eTable 1.

As a diet that broadly reflects dietary guidelines that do not take environmental impact into consideration, we also investigated the Alternate Healthy Eating Index (AHEI)-2010 score, as described in detail by Chiuve et al.^11^ Briefly, the score was based on foods and nutrients associated with chronic disease risk. The score included 11 components: vegetables, fruit, whole grains, sugar-sweetened beverages and fruit juice, nuts and legumes, red and processed meat, *trans* fat, long-chain n-3 fats, polyunsaturated fat, sodium, and alcohol. The score ranged from 0-110 points and was calculated as the sum of the relative adherence to each component (eTable 2).

### Stroke ascertainment

Incident cases of stroke were identified by linkage of each participant’s civil registration number to the Danish National Patient Registry. This registry covers discharge records from all Danish hospitals since 1977. Stroke was defined as the first hospital discharge diagnosis according to the International Classification of Disease (ICD-8 codes: 430, 431, 433, 434, 436.01 or 436.90; ICD-10 codes: I60, I61, I63 or I64). All registry-identified cases before 30 November 2009 were verified by review of medical records and/or hospital discharge letters.^12^

Stroke cases were classified as: ischemic stroke, intracerebral hemorrhage, subarachnoid hemorrhage or unspecified if we were unable to identify the subtype.

### Assessment of covariates

The lifestyle questionnaire assessed highest attained level of education, current smoking status (including type and amount), physical activity during summer and winter, and history of hypertension, hypercholesterolemia or diabetes as previous diagnosis or according to current use of relevant medication. History of myocardial infarction was identified using the Danish National Patient Registry. Alcohol intake was estimated from the FFQ and based on questions concerning type and frequency of consumption of alcoholic beverages. At the study center standardized measures of height, weight and waist circumference were conducted by trained personnel. Body mass index (BMI) was calculated as weight in kilograms divided by height in meters squared (kg/m^2^).

### Statistical analysis

Statistical analyses were conducted using Stata v16.1 (StataCorp, LLC 2019, College Station, TX). Baseline characteristics were reported using standard statistical summary measures.

Cox proportional hazards models were used to estimate the hazard ratios (HR) with corresponding 95% CIs for the association between adherence to the EAT-Lancet score and risk of stroke and subtypes. Age was the underlying time-scale. Follow-up was counted from entry into the study until censoring, which was incidence of stroke, death due to other causes, emigration or 30 November 2009; whichever came first. Analyses were repeated to assess the association between the AHEI and risk of stroke and subtypes. The proportional hazards assumptions were evaluated using log-log plots and no violations were observed.

The EAT-Lancet diet score was categorized into five groups based on the distribution of the data (0-7, 8, 9, 10 and 11-14 points). In supplementary analyses the score was also modelled using restricted cubic splines to assess potential non-linearity. To estimate risk differences (RD) we used the pseudo-observation method with time in the study as the underlying time-scale and death due to other causes as competing event. We estimated the 15 year risk of stroke.

We investigated several multivariable models. Model 1a, a crude model: adjusted for age (as underlying time scale), tertiles of baseline date of inclusion and age at inclusion (as strata). Model 1b, including potential confounders: further adjusted for education, smoking status, physical activity, alcohol intake and, in women, use of hormone replacement therapy. Model 2, including potential mediators: further adjusted for BMI, waist circumference, history of hypertension, hypercholesterolemia, diabetes and myocardial infarction.

In secondary analyses, we investigated effect modification on the absolute (RD) scale to assess whether certain subgroups may experience greater benefits than others when transitioning to a plant-based diet. We focused on the absolute scale because this is most relevant in terms of public health impact. We investigated subgroups by age, sex and BMI. The analysis of stroke subtypes was also stratified by sex to investigate whether the patterns of associations on the relative scale were different in men and women.

As a sensitivity analysis we excluded those at high risk at baseline, meaning those with previous myocardial infarction and/or diabetes as these conditions may lead to changed dietary intake and hence reverse causation. Because the cut off for dietary fiber in the component concerning grain intake in the EAT-Lancet diet score was low in relation to this Danish cohort, we set it to ≥ 25 g/day for women and ≥ 35 g/day for men, as this is the official recommended intake of dietary fiber in the Nordic countries.^13^

### Data availability

Data described in this article may be made available upon request pending on application to and approval by the Danish Cancer Society. Code is available upon request to the corresponding author.

## Results

Those with the greatest adherence to the EAT-Lancet diet score (11-14 points) were generally younger, more likely to be women and physically active, had longer education and had a lower BMI and waist circumference than those with the lowest level of adherence (0-7 points) (Table 1). They were, however, more likely to have a history of hypertension or hypercholesterolemia. Those who developed stroke were older at baseline, more likely men, had a shorter education, smoked, were physically inactive, had a larger waist circumference and had a history of hypertension, hypercholesterolemia or diabetes compared with the entire cohort (eTable 3). Compared with the other stroke subtypes, those who developed subarachnoid hemorrhage were more likely to be younger (eTable 4). They were also more likely to be female, physically inactive, have shorter education and smoke, and less likely to have a history of hypertension or hypercholesterolemia (eTable 3). Of the 2253 participants that developed stroke over the follow up period (median follow up time=15 years), 13 participants had an unknown cause of stroke and were not included in analyses of subtypes.

**Table 1.** Baseline characteristics of participants in the Danish Diet, Cancer and Health cohort across EAT-Lancet diet score groups^1^

| Characteristics | Cohort<br>n=55,016 |  |  | 0-7 points<br>n=6318 |  |  | 8 points<br>n=12,189 |  |  | 9 points<br>n=16,441 |  |  | 10 points<br>n=12,631 |  |  | 11-14 points<br>n=7437 |  |  |
| --- | --- | --- | --- | --- | --- | --- | --- | --- | --- | --- | --- | --- | --- | --- | --- | --- | --- | --- |
|  | Median<br>n | p10<br>% | p90 | Median<br>n | p10<br>% | p90 | Median<br>n | p10<br>% | p90 | Median<br>n | p10<br>% | p90 | Median<br>n | p10<br>% | p90 | Median<br>n | p10<br>% | p90 |
| **EAT-Lancet diet score** | 9 | 7 | 11 | 7.0 | 6.0 | 7.0 | 8.0 | 8.0 | 8.0 | 9.0 | 9.0 | 9.0 | 10.0 | 10.0 | 10.0 | 11.0 | 11.0 | 12.0 |
| **AHEI diet score** | 50 | 37 | 64 | 41.7 | 30.8 | 53.3 | 45.6 | 34.3 | 57.1 | 49.5 | 38.0 | 61.2 | 54.1 | 42.4 | 65.3 | 59.6 | 47.8 | 71.5 |
| **Age (years)** | 56 | 51 | 63 | 56.0 | 51.0 | 63.0 | 56.0 | 51.0 | 63.0 | 56.0 | 51.0 | 63.0 | 55.0 | 51.0 | 62.0 | 55.0 | 51.0 | 62.0 |
| **Men (%)** | 26208 | 48 |  | 4376 | 69 |  | 7202 | 59 |  | 8060 | 49 |  | 4671 | 37 |  | 1899 | 26 |  |
| **<9 years education (%)** | 8125 | 15 |  | 1173 | 19 |  | 2029 | 17 |  | 2471 | 15 |  | 1656 | 13 |  | 796 | 11 |  |
| **Current smoker (%)** | 19735 | 36 |  | 2938 | 47 |  | 4972 | 41 |  | 6002 | 37 |  | 3949 | 31 |  | 1874 | 25 |  |
| **Alcohol intake (g/d)** | 13 | 2 | 48 | 15.1 | 2.2 | 57.0 | 14.4 | 2.0 | 55.4 | 13.7 | 2.0 | 49.0 | 12.8 | 1.9 | 44.3 | 11.8 | 1.7 | 40.1 |
| **<30 min/day physical activity (%)** | 33257 | 60 |  | 3962 | 63 |  | 7628 | 63 |  | 10,149 | 62 |  | 7457 | 59 |  | 4061 | 55 |  |
| **Current use of HRT in women (%)** | 8667 | 30 |  | 556 | 29 |  | 1576 | 32 |  | 2493 | 30 |  | 2408 | 30 |  | 1634 | 30 |  |
| **Body mass index (kg/m<sup>2</sup>)** | 26 | 21 | 31 | 26.0 | 21.7 | 31.4 | 25.7 | 21.6 | 31.2 | 25.5 | 21.5 | 31.2 | 25.4 | 21.3 | 31.1 | 25.0 | 21.2 | 30.8 |
| **Waist circumference in men (cm)** | 95 | 84 | 109 | 96 | 84 | 109 | 95 | 84 | 109 | 95 | 84 | 109 | 95 | 84 | 108 | 94 | 83 | 107 |
| **Waist circumference in women (cm)** | 80 | 69 | 97 | 82 | 70 | 99 | 8 | 70 | 98 | 81 | 70 | 98 | 80 | 69 | 97 | 79 | 69 | 96 |
| **History of hypertension (%)** | 8773 | 16 |  | 848 | 13 |  | 1895 | 16 |  | 2709 | 16 |  | 2056 | 16 |  | 1265 | 17 |  |
| **History of hypercholesterolemia (%)** | 4043 | 7 |  | 373 | 6 |  | 840 | 7 |  | 1245 | 8 |  | 1010 | 8 |  | 575 | 8 |  |
| **History of diabetes (%)** | 1107 | 2 |  | 79 | 1 |  | 191 | 2 |  | 319 | 2 |  | 288 | 2 |  | 230 | 3 |  |
| **History of myocardial infarction (%)** | 831 | 2 |  | 115 | 2 |  | 190 | 2 |  | 281 | 2 |  | 162 | 1 |  | 83 | 1 |  |

<sup>1</sup>All medians and percentiles are pseudo-medians and pseudo-percentiles meaning that they represent the average of the 5 surrounding values in order to comply with European data protection regulations. AHEI: Alternate Healthy Eating Index, HRT: hormone replacement therapy.

Those with the greatest adherence to the EAT-Lancet diet also had greater adherence to the AHEI (Table 1). Of those with the poorest adherence to the EAT-Lancet diet (0-7 point) 74% were within the two lowest AHEI groups (13-46 points) and 77% of those with the greatest adherence to the EAT-Lancet diet (11-14 points) were within the two highest AHEI groups (53-110 points) (eFigure 5). One of the main differences in dietary intake between the extreme intake groups of the two diets were intake of whole grains, which was highest among those with the greatest adherence to the AHEI and indifferent between the EAT-Lancet diet groups (Table 2). The contrast in differences between extremes was greater for intake of eggs and dairy products, as participants having a low EAT-Lancet diet score reported more of these food groups than participants having a low score on the AHEI.

**Table 2.** Dietary intake (median, 10^th^ and 90^th^ percentiles^1^) in the highest and lowest adherence groups of the EAT-Lancet diet and the AHEI

| Characteristics | EAT-Lancet diet score |  |  |  |  |  | AHEI score |  |  |  |  |  |
| --- | --- | --- | --- | --- | --- | --- | --- | --- | --- | --- | --- | --- |
|  | 0-7 points<br>n=6318 |  |  | 11-14 points<br>n=7437 |  |  | 0-40 points<br>n=10,919 |  |  | 59-110 points<br>n=11,054 |  |  |
|  | Median | p10 | p90 | Median | p10 | p90 | Median | p10 | p90 | Median | p10 | p90 |
| EAT-Lancet diet score, points | 7 | 6 | 7 | 11 | 11 | 12 | 8 | 7 | 10 | 10 | 9 | 11 |
| AHEI diet score, points | 42 | 31 | 53 | 60 | 48 | 72 | 37 | 30 | 40 | 64 | 60 | 72 |
| Whole grains, g/d | 132 | 65 | 237 | 128 | 64 | 223 | 119 | 63 | 218 | 134 | 70 | 227 |
| Refined grains, g/d | 59 | 24 | 123 | 37 | 17 | 81 | 62 | 26 | 128 | 35 | 16 | 72 |
| Potatoes, g/d | 169 | 109 | 315 | 80 | 39 | 159 | 156 | 74 | 299 | 103 | 45 | 212 |
| Vegetables, g/d | 129 | 57 | 213 | 229 | 108 | 358 | 123 | 52 | 233 | 212 | 101 | 381 |
| Fruit, g/d | 79 | 24 | 285 | 201 | 111 | 447 | 88 | 21 | 230 | 229 | 80 | 496 |
| Red and processed meat/beef, lamb, pork, g/d | 139 | 76 | 236 | 79 | 30 | 142 | 152 | 98 | 239 | 69 | 33 | 112 |
| Chicken and other poultry, g/d | 19 | 6 | 62 | 18 | 5 | 41 | 18 | 5 | 40 | 18 | 5 | 48 |
| Eggs, g/d | 40 | 19 | 71 | 14 | 6 | 25 | 27 | 11 | 61 | 18 | 7 | 46 |
| Fish, g/d | 44 | 19 | 107 | 35 | 15 | 66 | 41 | 14 | 82 | 37 | 18 | 72 |
| Dairy products, g/day | 591 | 127 | 1061 | 242 | 60 | 454 | 283 | 66 | 833 | 284 | 73 | 729 |
| Peanuts and tree nuts, g/d | 1 | 0 | 4 | 1 | 0 | 4 | 1 | 0 | 4 | 1 | 0 | 4 |
| Sugar-sweetened beverages, g/d | 18 | 1 | 201 | 4 | 0 | 29 | 30 | 1 | 202 | 4 | 0 | 18 |
| Added sugar/all sweeteners, g/d | 45 | 23 | 91 | 21 | 8 | 34 | 45 | 17 | 97 | 22 | 9 | 45 |
| Polyunsaturated fat, E% | 5 | 4 | 7 | 6 | 4 | 8 | 5 | 4 | 7 | 6 | 4 | 8 |
| Dietary fiber, g/d | 20 | 13 | 30 | 22 | 14 | 31 | 19 | 12 | 27 | 23 | 15 | 34 |

<sup>1</sup>All medians and percentiles are pseudo medians and pseudo percentiles meaning that they represent the average of the 5 surrounding values in order to comply with European data protection regulations. AHEI: Alternate Healthy Eating Index.

After adjustment for potential confounders, the risk of total stroke was lower, although the CIs were wide and overlapped no association, among those with the greatest adherence to the EAT-Lancet diet compared to those with the poorest adherence (HR 0.91; 95% CI 0.76, 1.09; RD% - 0.9; 95% CI -2.0, 0.2%; Figure 1). A lower risk of total stroke was observed for adherence to the AHEI (HR 0.75; 95% CI 0.64, 0.87; RD% -1.34; 95% CI -2.26, -0.43%; Figure 1). The patterns of associations were similar after further adjustment for potential mediators on the relative and absolute scale for the EAT-Lancet diet score (eFigure 2-3) and AHEI (eFigure 4-5). When modelling the EAT-Lancet diet score using restricted cubic splines, the curve suggested a threshold of no association until 9 points and then a linear decrease in risk (eFigure 6). A linear pattern was observed for the AHEI (eFigure 6).

**Figure 1.**
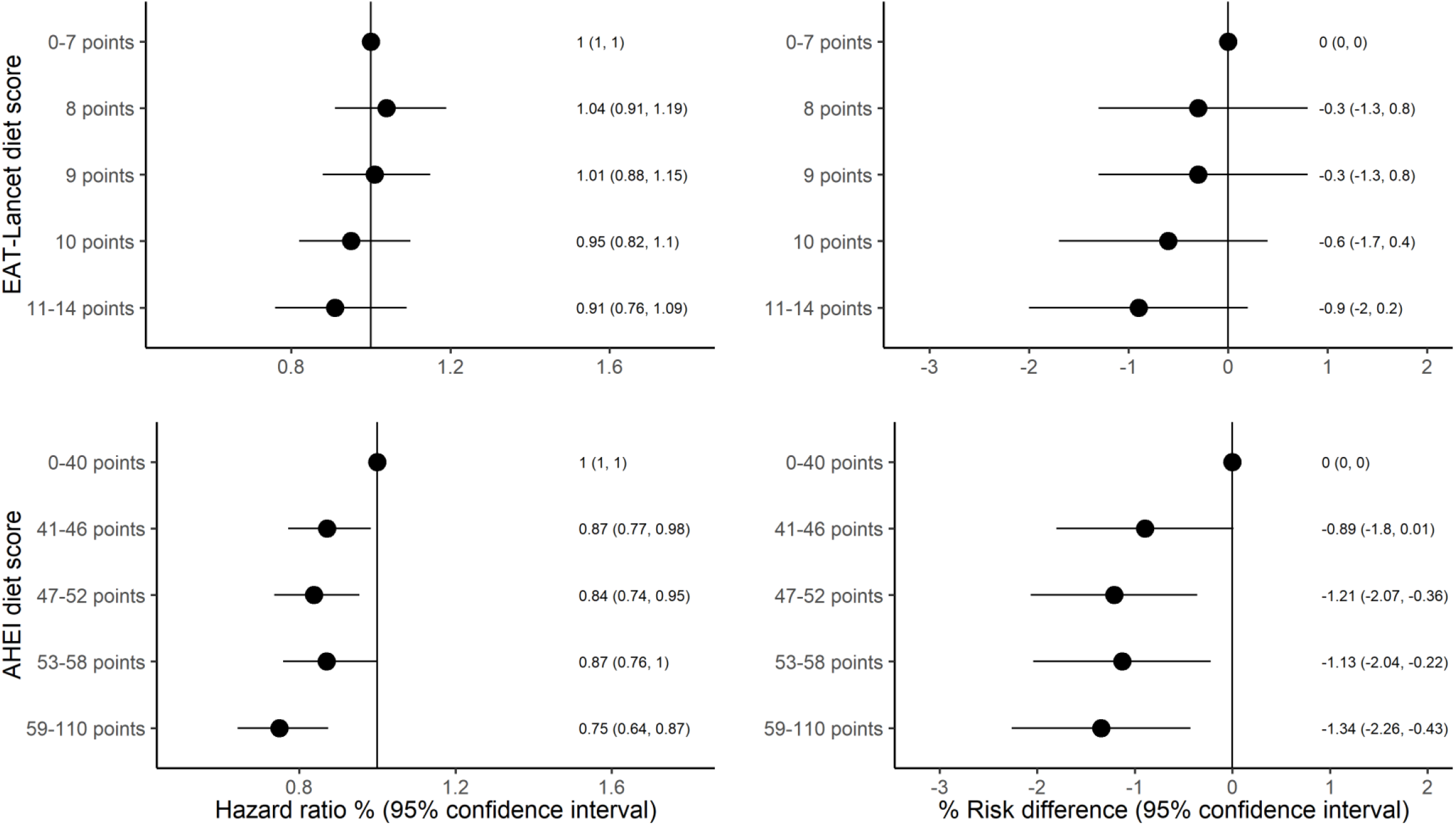
Diet scores and stroke risk. Association between adherence to the EAT-Lancet diet score or the Alternate Healthy Eating Index (AHEI) diet score and risk of stroke in middle aged adults. Hazard ratio adjusted for age (underlying time scale), sex, date of inclusion and age at inclusion (as strata), education, smoking status, physical activity, alcohol intake and hormone replacement therapy. Risk difference adjusted for age, sex, education, smoking status, physical activity, alcohol intake and hormone replacement therapy. N = 55,016, N cases = 2253 (2251 cases after 15 years of follow-up).

For stroke subtypes, we found that adherence to the EAT-Lancet diet was associated with a lower risk of subarachnoid hemorrhage (HR 0.30; 95% CI 0.12, 0.73) but not ischemic stroke or intracerebral hemorrhage (Figure 2). In contrast, the AHEI was associated with a lower risk of ischemic stroke (HR 0.76; 95% CI 0.64, 0.90) and intracerebral hemorrhage (HR 0.58; 95% CI 0.36, 0.93) but not subarachnoid hemorrhage (Figure 2). For the EAT-Lancet score, after we stratified by sex, a lower risk of subarachnoid hemorrhage was observed in women (HR 0.26, 95% CI 0.06, 0.81, n cases=77) but not statistically significant in men (HR 0.53, 95% CI 0.11, 2.49, n cases=38) (eFigure 7). Note, that the HRs are not directly comparable between men and women as they have different baseline hazards.

In secondary analyses, the absolute risk difference of total stroke comparing the greatest to poorest adherence to the EAT-Lancet diet was lower among those aged <57 years than in those aged ≥ 57 years at baseline, in men than women and among those with a BMI<25 than those with a BMI ≥ 25 kg/m^2^ (Figure 3).

**Figure 2.**
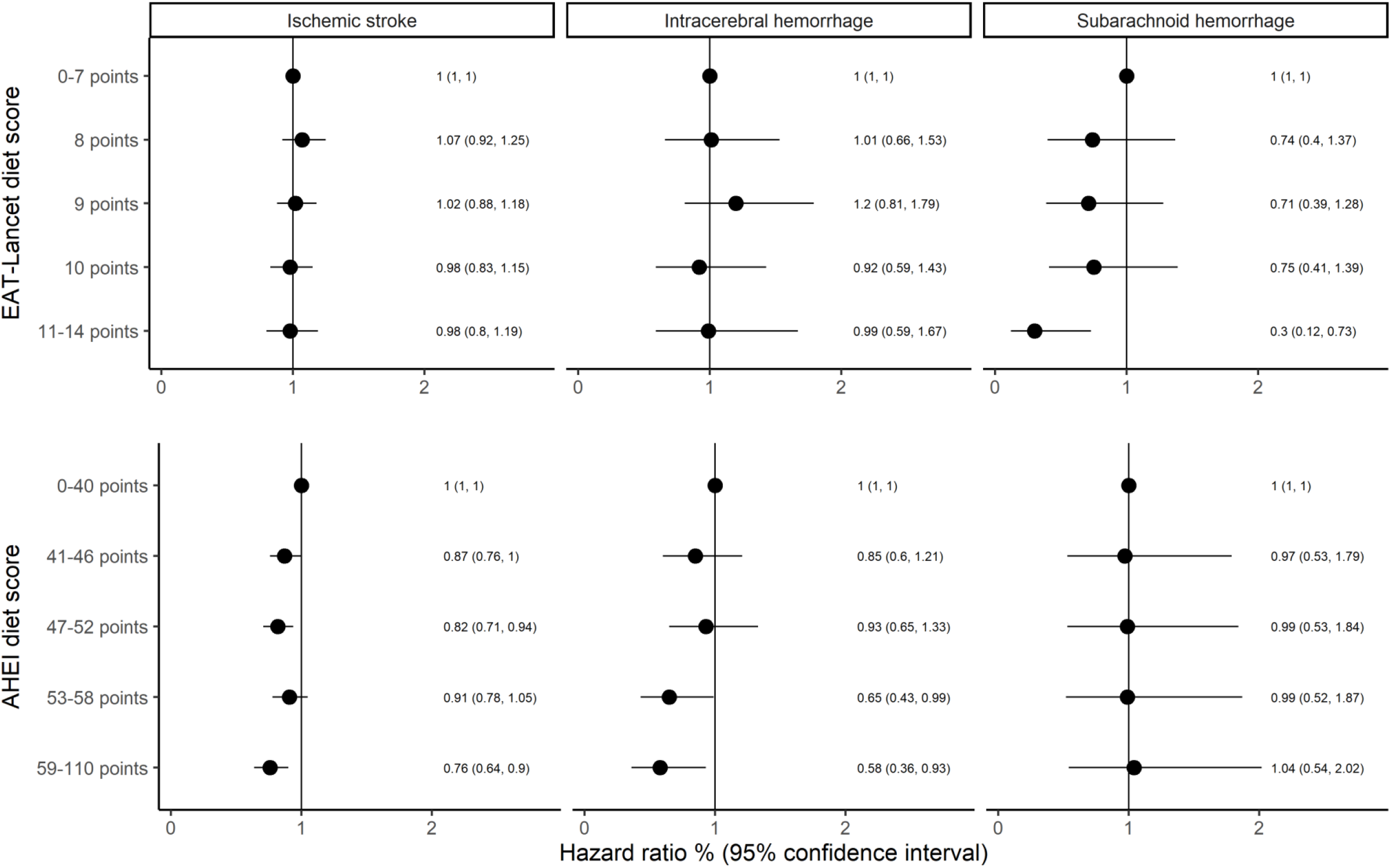
Diet scores and stroke subtype risk. Association between adherence to the EAT-Lancet diet score or the Alternate Healthy Eating Index (AHEI) score and risk of stroke subtypes in middle aged adults. Adjusted for age (underlying time scale), sex, date of inclusion and age at inclusion (as strata), education, smoking status, physical activity, alcohol intake and hormone replacement therapy. N = 55,016, N intracerebral hemorrhage cases = 267, N ischemic stroke cases = 1858, N subarachnoid hemorrhage cases = 115.

**Figure 3.**
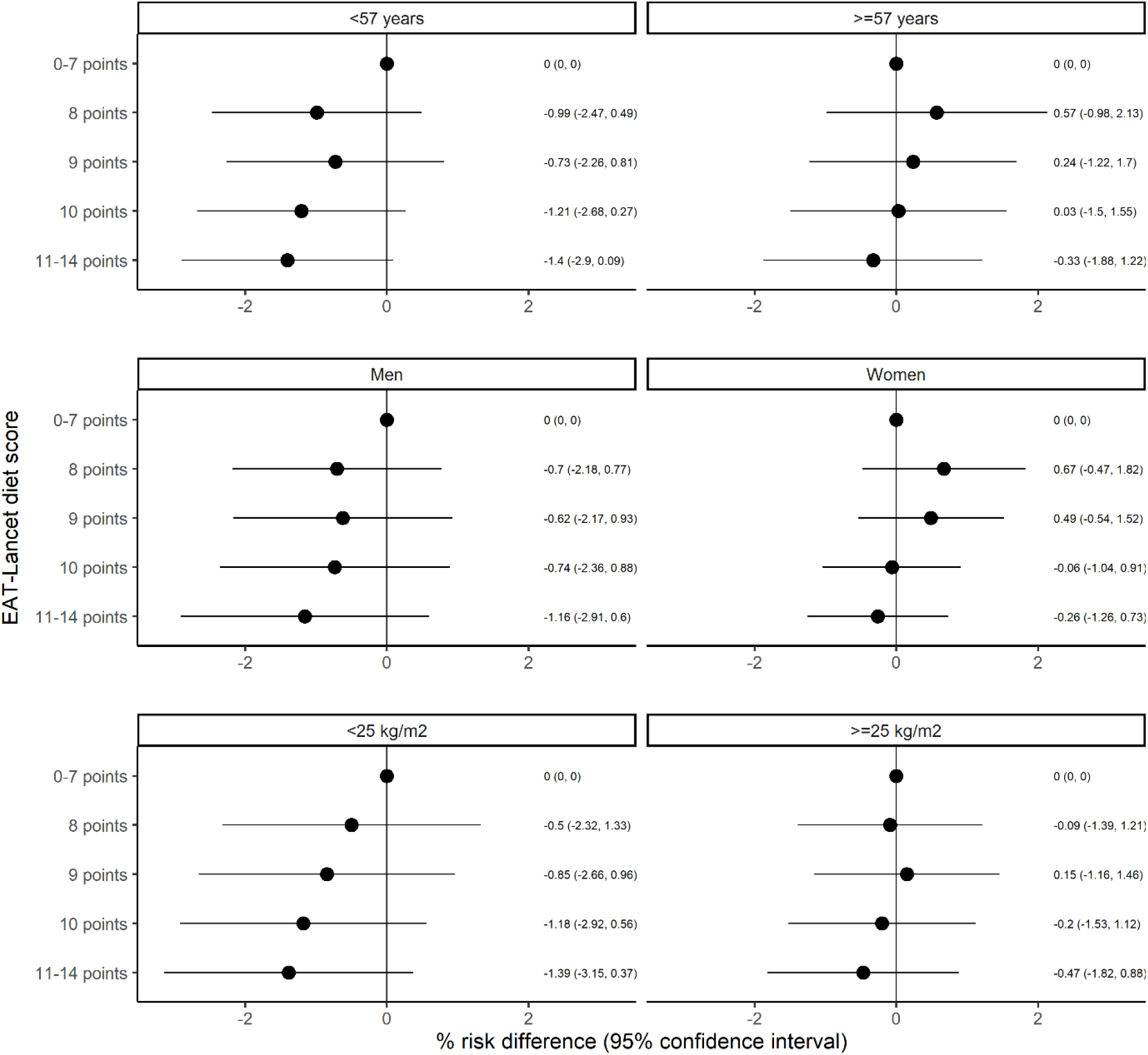
EAT-Lancet diet and risk of stroke stratified by age, sex and BMI. Association between adherence to the EAT-Lancet diet score and risk of stroke in subgroups by age, sex and BMI. Risk differences were adjusted for age, sex, education, smoking status, physical activity, alcohol intake and hormone replacement therapy. For age: <57 years: N = 30,630, N cases = 887. >=57 years: N = 24,386, N cases = 1366. P for interaction=0.79. For sex: Men: N = 26,208, N cases = 1354. Women: N = 28,808, N cases = 897. P for interaction=0.62. For BMI: BMI <25 kg/m^2^: N = 24,131, N cases = 854. BMI >=25 kg/m^2^: N = 30,885, N cases = 1397. P for interaction=0.13.

In sensitivity analyses, we found similar patterns as those in the main analysis after we excluded those with a history of diabetes or myocardial infarction before study start (eFigure 8). When the threshold for dietary fiber intake was increased for the EAT-Lancet diet component concerning grains, only 14% (n=7806) of the participants adhered to the component, compared with 99.7% (n=54,849) at the original threshold. With the updated score, we observed a lower risk of stroke for those with the greatest adherence compared to those with the lowest adherence to the EAT-Lancet diet, both on the relative scale (HR 0.78, 95% CI 0.60, 1.01, Model 2, HR 0.75, 95% CI 0.58, 0.97, Model 3) and additive scale (RD% -1.05; 95% CI -1.90, -0.20%, Model 2, RD% -1.11; 95% CI -1.96, -0.26%, Model 3).

## Discussion

In this cohort of middle aged Danish adults we found that adherence to the EAT-Lancet diet was associated with a lower risk of stroke, but with some uncertainty and mainly driven by a lower risk of subarachnoid hemorrhage. Greater adherence to the AHEI was more clearly associated with a lower risk of total stroke, which, in contrast to the EAT-Lancet diet, seemed to be related to a lower risk of ischemic stroke and intracerebral hemorrhage, suggesting that each dietary pattern had a distinct association with specific stroke subtypes.

One of the strengths of this study is that it used register identified stroke cases that were subsequently validated and subtyped using medical records.^12^ The distribution of stroke subtypes was representative of typical Western populations with 83% ischemic strokes, 12% intracerebral hemorrhages and 5% subarachnoid hemorrhages.^14^ Only 13 cases were not classified according to stroke subtype. Because we used registers to identify stroke cases, we also ensured that few participants were lost to follow up. In total only 0.4% (n=212) emigrated during the follow up period. To classify participants according to the different dietary pattern scores we used data derived from a FFQ. Although the FFQ was able to rank participants into similar quintiles of macronutrient intake as determined by two 7 day food records in our validation study, there is still a risk of misclassification. Our prospective design ensures that participants likely were unaware of their future risk of stroke, which would, on average, bias our estimates towards the null. In the same vein, we only used a single measure of dietary intake, which weakens inference regarding long term dietary intake, as participants could have changed their diets. After excluding participants at high risk at baseline, those with previous myocardial infarction or diabetes, we observed similar patterns of associations. Given our observational study design, we cannot exclude the possibility of residual confounding. We adjusted for many known social and behavioral risk factors for stroke that are also likely to be related to food intake, but potential confounders such as sleep^15^ and stress^16^ were not taken into account.

The EAT-Lancet diet was constructed by weighing health and environmental considerations^2^ whereas the AHEI was constructed based on the relation between individual dietary components and their association with chronic diseases, particularly cardiovascular diseases^11^. The difference in the associations with stroke and stroke subtypes for the two dietary patterns investigated may therefore be explained by the food components in each diet score. The lower risk observed with greater adherence to the AHEI seemed to be driven by the most frequent stroke subtypes, ischemic stroke and intracerebral hemorrhage. In contrast, greater adherence to the EAT-Lancet diet was mainly related to a lower risk of subarachnoid hemorrhage. Although the two diets classified similar participants in the highest and lowest intake groups, there were some differences between the diet scores. One difference was that the AHEI focused more on specific dietary factors related to lower risk of atherosclerotic related cardiovascular diseases such as whole grains, omega-3 fatty acids and sodium.^5^ In particular, in our Danish cohort with a higher fiber intake, almost all participants adhered to the EAT-Lancet grain component. When we, in a sensitivity analysis, increased the fiber threshold to current Nordic fiber recommendations, we found a lower risk of stroke similar to that of the AHEI. Previous studies have also showed that intake of dietary fiber was associated with lower risk of ischemic, but not hemorrhagic strokes.^17^ Another difference was the construction of the diet scores. The EAT-Lancet diet was scored based on adherence vs no adherence whereas the AHEI gave a relative score for gradual adherence to the diet, providing a greater differentiation between those with high and low adherence. This was evident from the spline analysis, as the risk of stroke was gradually lower with greater adherence, whereas the EAT-Lancet diet score showed a threshold pattern. Hence, the scoring of the EAT-Lancet diet may result in poorer differentiation between the non-extreme intake groups, whereas the extremes of the diet score would be similar.

The potential beneficial association between greater adherence to the EAT-Lancet diet and risk of total stroke may be due to higher intake of vegetables, fruits and lower intakes of red and processed meat, which has also been observed in other cohort studies, particularly for ischemic stroke risk.^17^ Diets with higher levels of fruits and vegetables have been related to lower blood pressure in several trials^18^, likely through their potassium content as well as displacement of processed foods high in sodium. Red meat contains potentially risk increasing nutrients such as saturated fat, cholesterol, and heme iron and sodium for processed meat^5,19^. The relation between greater adherence to the EAT-Lancet diet and lower risk of subarachnoid hemorrhage may be due to lower intakes of poultry, eggs and dairy products, all foods that were not directly part of the AHEI. In a similar European cohort, a higher intake of eggs was associated with a higher risk of hemorrhagic stroke.^17^ A meta-analysis of cohorts suggested there to be a J shaped association between higher intake of eggs (and a concomitant lower intake of other foods) and risk of total stroke^20^. Eggs contain high amounts of cholesterol and intakes of >40-50 g/day of eggs may be associated with a higher risk of stroke^20^. They are, however, also an indicator of a diet high in eggs, which, in some populations, may also explain the higher risk observed with greater intakes. Dairy product intake was associated with a lower risk of total stroke in a meta analysis of cohort studies, potentially due to the role of calcium, potassium and bioactive peptides related to lower blood pressure.^21^ Low intake in our Danish cohort was around 240 g/day, which is consistent with a lower risk of hemorrhagic stroke and, in the dose-response meta analysis, above which higher risks of hemorrhagic stroke was observed for milk intake, potentially due to higher intake of saturated fat.^21^ A meta analysis of cohort studies suggested that intake of poultry was not associated with risk of total stroke, but the evidence was inconsistent and depended on the region investigated.^22^ In general, few of these studies investigated subtypes of stroke and those that did, investigated ischemic and hemorrhagic stroke in broad groupings.

Vegetarians in the EPIC-Oxford cohort had a higher risk of total stroke and hemorrhagic stroke compared with meat eaters.^3^ This raised the question of whether or not adherence to a plant-based diet, like the EAT-Lancet diet, could have unintended health consequences. In our study of middle-aged Danish adults, of which the vast majority consumed meat, we could not confirm this concern. In fact, our results, if anything, indicated the opposite. Namely, that adherence to the EAT-Lancet diet was associated with a lower risk of stroke, particularly subarachnoid hemorrhage. These two studies have substantial differences in diets, populations and distribution of stroke subtypes investigated. Compared to those with the greatest adherence to the EAT-Lancet diet in this study, the vegetarians in EPIC-Oxford had a higher intake of fruit, vegetables, nuts and legumes and a lower intake of meat. It is unlikely that these differences in dietary intake explain the observed differences, given that intake of fruit and vegetables have been associated with a lower risk of ischemic stroke, not hemorrhagic stroke, whereas intake of nuts and legumes was not associated with ischemic or hemorrhagic stroke in a large European cohort.^17^ The vegetarians and vegans in the EPIC-Oxford cohort have likely adhered to their diet for many years as many upon recruitment were members of a vegetarian society.^3^ Consequently, their nutritional status may be different from our Danish general population, specifically with regards to levels of vitamin B12,^23^ vitamin D^24^ and omega-3 fatty acids, ^25^ all suggested to play a role in the development of stroke, although the evidence is not convincing.^26^ The distribution of stroke cases in the EPIC-Oxford study was also different compared with typical Western patterns, with 48% ischemic strokes, 28% hemorrhagic stroke and 24% strokes of unknown etiology. The EPIC-Oxford cohort included a younger age group at recruitment and, as was also evident in our study, those who developed subarachnoid hemorrhage were younger, although there was no information on subtypes of hemorrhagic stroke in the EPIC-Oxford study. In two Taiwanese cohorts including vegetarians, a lower risk of both ischemic and hemorrhagic stroke was found compared with nonvegetarians.^27^ In three US cohorts of health professionals and adherence to a healthy plant-based diet was associated with a lower risk of total stroke and uncertainty around the estimates concerning ischemic and hemorrhagic strokes.^4^ All in all, this suggests that the EPIC-Oxford results should be interpreted with caution and may not be directly generalizable to other populations.

In our subgroup analyses, we did not find a clear public health benefit of following the EAT-Lancet diet depending on age, sex or BMI, which may be because the EAT-Lancet diet was mainly associated with a lower risk of subarachnoid hemorrhage, the stroke subtypes with the fewest cases. For adherence to the AHEI, our analysis indicated that 1.34% of the cases could have been prevented if participants with the poorest adherence shifted to the greatest adherence. When the fiber threshold was increased for adherence to the EAT-Lancet diet, 1.05% of the cases could have been prevented. Thus, to enhance the public health benefit of adhering to the EAT-Lancet diet, more emphasis could be placed on dietary fiber. It is important to keep the study context in mind and note that this Danish population generally had high intakes of red meat and dairy. This population also had a very low intake of nuts and legumes. Thus, these findings may not be directly transferrable to contexts with different dietary intakes.

In conclusion, these results suggest that adherence to the EAT-Lancet diet was associated with a lower risk of stroke, although not statistically significant, and particularly associated with a lower risk of subarachnoid stroke. Adherence to the AHEI was associated with a lower risk of total stroke, mainly ischemic and intracerebral hemorrhage. Future studies should further investigate the potential differential role of dietary patterns on stroke subtype risk and the long term health consequences of shifting from a meat-based diet to a plant-based diet.

## Supporting information

eTable

## Data Availability

Data described in this article may be made available upon request pending on application to and approval by the Danish Cancer Society.

https://www.cancer.dk/research/dcrc-research/diet-cancer-and-health/

## Acknowledgements

We thank the Danish Cancer Society, the staff at the Danish Diet, Cancer and Health study for collection of the data and the study participants for their contribution to the study.

## Appendix 1 Authors

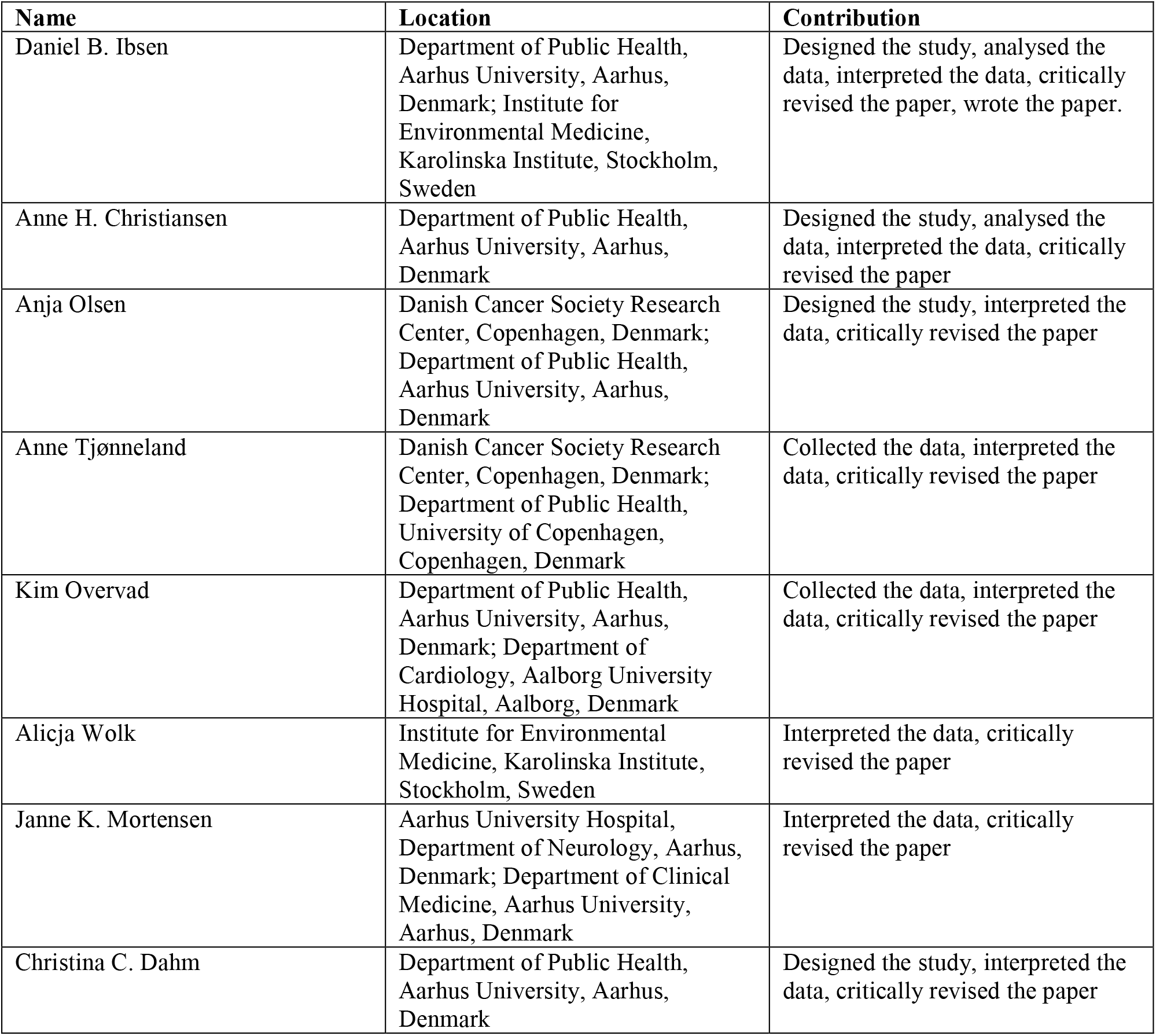

