## Supplementary material for "Adherence to the EAT-Lancet diet and risk of stroke and stroke subtypes: A Danish cohort study": eTable

### Supplemental Tables

**eTable 1** Construction of the EAT-Lancet diet score in the Danish Diet, Cancer and Health cohort

| Dietary components | Original EAT-Lancet macronutrient intake (possible range)[1] | Criteria for scoring 1 point[2] based on the EAT-Lancet reference diet[3] | Food items based on the available foods in the food frequency questionnaire in the Danish Diet, Cancer and Health cohort | n (%) participants scoring 1 point for each dietary component (n=55,016) |
|---|---|---|---|---|
| **Grains including whole grains** | | | | |
| 1. Rice, wheat, corn and other | 232 (total grains 0-60% of energy) g/day | ≤464g/day and whole grain fiber >5g/day | Whole grains cereals, refined grain cereals | 54,849 (100%) |
| **Tubers and starchy vegetables** | | | | |
| 2. Potatoes and casava | 50 (0-100) g/day | ≤100g/day | Potatoes, fatty potatoes, | 18,820 (34%) |
| **Vegetables** | | | | |
| 3. All vegetables | 300 (200-600) g/day | ≥200g/day | Leafy vegetables, fruiting vegetables, cabbages, mushrooms, onion/garlic, stalk vegetables, other root vegetables | 18,280 (33%) |
| **Fruits** | | | | |
| 4. All fruits | 200 (100-300) g/day | ≥100g/day | Citrus fruits, other fruits | 36,009 (65%) |
| **Dairy products** | | | | |
| 5. Whole milk or derivative equivalents | 250 (0-500) g/day | ≤500g/day | Skimmed milk, semi-skimmed milk, whole milk, buttermilk, cheese, whole-fat fermented, low-fat fermented | 38,911 (71%) |
| **Protein sources** | | | | |
| 6. Beef, lamb, pork | 14 (0-28) g/day | ≤28g/day | Red meat, processed meat | 1088 (2%) |
| 7. Chicken and other poultry | 29 (0-58) g/day | ≤58g/day | Poultry | 52,181 (95%) |
| 8. Eggs | 13 (0-25) g/day | ≤25g/day | Eggs | 31,458 (57%) |
| 9. Fish | 28 (0-100) g/day | ≤100g/day | Fish/lean/fresh, fish/lean/processed, fish/med fat/fresh, fish/med fat/processed, fish/fatty/fresh, fish/fatty/processed | 53,092 (97%) |
| **Legumes** | | | | |
| 10. Dry beans, lentils, peas | 50 (0-100) g/day | ≤100g/day | Legumes | 55,016 (100%) |
| 11. Soy foods | 25 (0-50) g/day | ≤50g/day | Soy | 55,016 (100%) |
| 12. Peanuts or tree nuts | 25 (0-75) g/day | ≥25g/day | Nuts | 215 (0%) |
| **Added fats** | | | | |

| | | | | |
|---|---|---|---|---|
| 13. Palm oil, unsaturated oils, dairy fats (incl. in milk), lard or tallow | Individual limits for palm oil, unsaturated fat, dairy fats, lard and tallow | Ratio of 0.8 for unsaturated:saturated fat intake | Saturated fatty acids, monounsaturated fatty acids, polyunsaturated fatty acids | 54,669 (99%) |
| **Added sugar** | | | | |
| 14. All sweeteners | 31 (0-31) g/day | ≤31g/day | Added sugar | 28,682 (52%) |

[1]From Willett et al.[e1]
[2]Each dietary component contributed 0 or 1 point resulting in a score ranging from 0 to 14 points.
[3]From Knuppel et al.[e2]

**eTable 2** Construction of the Alternate Healthy Eating Index-2010 score in the Danish Diet, Cancer and Health cohort based on Chiuve et al.[1]

| Dietary components | Unit | Criteria for scoring 0 point | Criteria for scoring 10 points | n (%) participants adhere[2] (n=55,016) |
|---|---|---|---|---|
| **Vegetables** | Servings/day; 1 serving=118 g vegetables (excl. potatoes) or 236.6 g green leafy vegetables | 0 | ≥5 | 126 (0.2%) |
| **Fruit** | Servings/day; 1 serving=1 piece of medium-sized fruit or 118 g berries | 0 | ≥4 | 3259 (5.9%) |
| **Whole grains** | g/day | | | 2622 (4.8%) |
| - men | | 0 | 90 | |
| - women | | 0 | 70 | |
| **Omega-3 fatty acids** | mg/day | 0 | 250 | 48,499 (88.2%) |
| **Polyunsaturated fat** | E% | ≤2 | ≥10 | 161 (0.3%) |
| **Nuts and legumes** | Servings/day; 1 serving=28 g or 15 ml peanut butter | 0 | ≥1 | 253 (0.5%) |
| **Sugar-sweetened beverages** | Servings/day; 1 serving=227 g | ≥1 | 0 | 295 (0.5%) |
| **Red and processed meat** | Servings/day; 1 serving=113 g red meat or 43 g processed meat | ≥1.5 | 0 | 217 (0.4%) |
| **Trans fat** | E% | ≥4 | ≤0.5 | 4914 (8.9%) |
| **Sodium** | mg/day | Highest fractile | Lowest fractile | 4910 (8.9%) |
| **Alcohol** | units/day; 1 unit=113 ml vine, 340 ml beer or 43 ml spirits | | | 23,891 (43.4%) |
| - men | | ≥3.5 | 0.5-2.0 | |
| - women | | ≥2.5 | 0.5-1.5 | |

[1]From Chiuve et al.[e3]

[2]Adherence defined as scoring 10 point for each dietary component

**eTable 3** Baseline characteristics of participants in the Danish Diet, Cancer and Health cohort across stroke subtypes[1]

| | Cohort n=55,016 | | | Total stroke n=2253 | | | Ischemic stroke n=1858 | | | Intralcerebral hemorrhage n=276 | | | Subarachnoid hemorrhage n=115 | | |
|---|---|---|---|---|---|---|---|---|---|---|---|---|---|---|---|
| **Characteristics** | **Median n** | **p10 %** | **p90** | **Median n** | **p10 %** | **p90** | **Median n** | **p10 %** | **p90** | **Median n** | **p10 %** | **p90** | **Median n** | **p10 %** | **p90** |
| **EAT-Lancet diet score** | 9 | 7 | 11 | 9 | 7 | 10 | 9 | 7 | 11 | 9 | 7 | 11 | 9 | 7 | 10 |
| **AHEI diet score** | 50 | 37 | 64 | 48 | 35 | 61 | 47 | 35 | 61 | 47 | 36 | 61 | 51 | 38 | 64 |
| **Age (years)** | 56 | 51 | 63 | 58 | 51 | 64 | 58 | 51 | 63 | 59 | 51 | 64 | 55 | 51 | 63 |
| **Men (%)** | 26208 | 48 | | 1356 | 60 | | 1148 | 62 | | 160 | 60 | | 38 | 33 | |
| **<9 years education (%)** | 8125 | 15 | | 398 | 18 | | 330 | 18 | | 45 | 17 | | 22 | 19 | |
| **Current smoker (%)** | 19735 | 36 | | 1125 | 50 | | 937 | 50 | | 117 | 44 | | 61 | 53 | |
| **Alcohol intake (g/d)** | 13 | 2 | 48 | 15 | 2 | 61 | 15 | 2 | 61 | 19 | 3 | 65 | 12 | 2 | 57 |
| **<30 min/day physical activity (%)** | 33257 | 60 | | 1480 | 66 | | 1208 | 65 | | 179 | 67 | | 83 | 72 | |
| **Current use of HRT in women (%)** | 8667 | 30 | | 301 | 34 | | 241 | 34 | | 36 | 34 | | 23 | 30 | |
| **Body mass index ($kg/m^2$)** | 26 | 21 | 31 | 26 | 22 | 32 | 26 | 22 | 32 | 26 | 21 | 32 | 25 | 20 | 30 |
| **Waist circumference in men (cm)** | 95 | 84 | 109 | 97 | 85 | 111 | 97 | 85 | 111 | 98 | 86 | 111 | 95 | 83 | 106 |
| **Waist circumference in women (cm)** | 80 | 69 | 97 | 82 | 70 | 102 | 82 | 70 | 102 | 78 | 68 | 100 | 81 | 70 | 96 |
| **History of hypertension (%)** | 8773 | 16 | | 624 | 28 | | 518 | 28 | | 78 | 29 | | 25 | 22 | |
| **History of hypercholesterolemia (%)** | 4043 | 7 | | 247 | 11 | | 195 | 10 | | 39 | 15 | | 11 | 10 | |
| **History of diabetes (%)** | 1107 | 2 | | 97 | 4 | | 86 | 5 | | 10 | 4 | | 0 | 0 | |
| **History of myocardial infarction (%)** | 831 | 2 | | 74 | 3 | | 63 | 3 | | 7 | 3 | | <5 | <5 | |

[1]All medians and percentiles are pseudo-medians and pseudo-percentiles meaning that they represent the average of the 5 surrounding values and groups with less than 5 individuals are masked in order to comply with European data protection regulations. AHEI: Alternate Healthy Eating Index, HRT: hormone replacement therapy.

**eTable 4** Cross-tabulation of stroke subtypes and age groups

| Stroke subtype, n (%) | Age group | | | | | |
|---|---|---|---|---|---|---|
| | **50-52 years** | **53-54 years** | **55-57 years** | **58-61 years** | **62-65 years** | **Total** |
| **Ischemic stroke** | 210 (11.3%) | 257 (13.8%) | 352 (18.9%) | 436 (23.5%) | 603 (32.5%) | 1858 |
| **Intracerebral hemorrhage** | 30 (11.2%) | 31 (11.6%) | 39 (14.6%) | 72 (27.0%) | 95 (35.6%) | 267 |
| **Subarachnoid hemorrhage** | 23 (20.0%) | 22 (19.1%) | 29 (25.2%) | 21 (18.3%) | 20 (17.4%) | 115 |

**eTable 5** Cross-tabulation of adherence groups of the EAT-Lancet diet and Alterative Healthy Eating Index (AHEI).

| EAT-Lancet group | AHEI group | | | | |
|---|---|---|---|---|---|
| | **13-40 points** | **41-46 points** | **47-52 points** | **53-58 points** | **59-110 points** |
| **0-7 points** | 3017 | 1646 | 952 | 506 | 197 |
| **8 points** | 3782 | 3207 | 2605 | 1769 | 826 |
| **9 points** | 2960 | 3737 | 3802 | 3502 | 2440 |
| **10 points** | 1003 | 1885 | 2638 | 3428 | 3677 |
| **11-14 points** | 157 | 526 | 1005 | 1835 | 3914 |

### Supplemental Figures

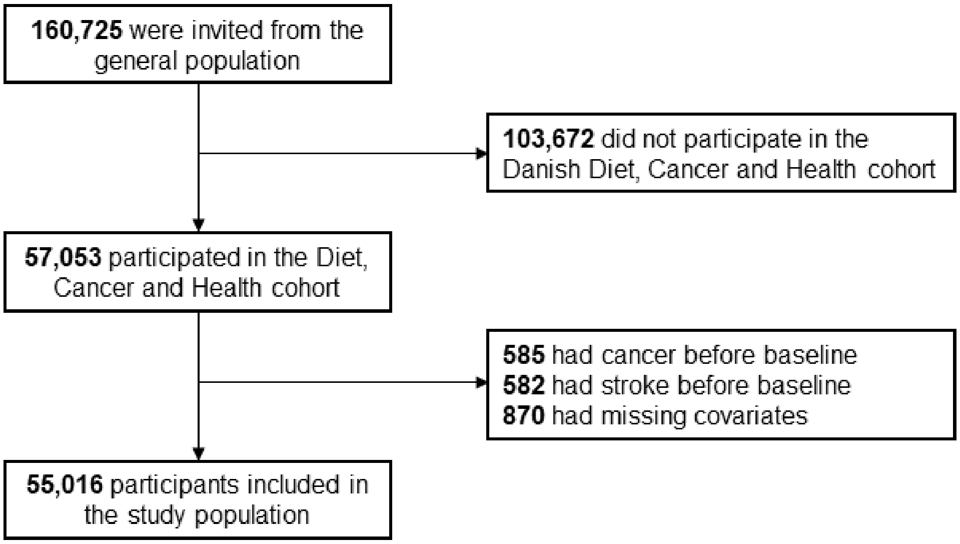


**eFigure 1** Flowchart of participants from the Danish Diet, Cancer and Health cohort included in this study.

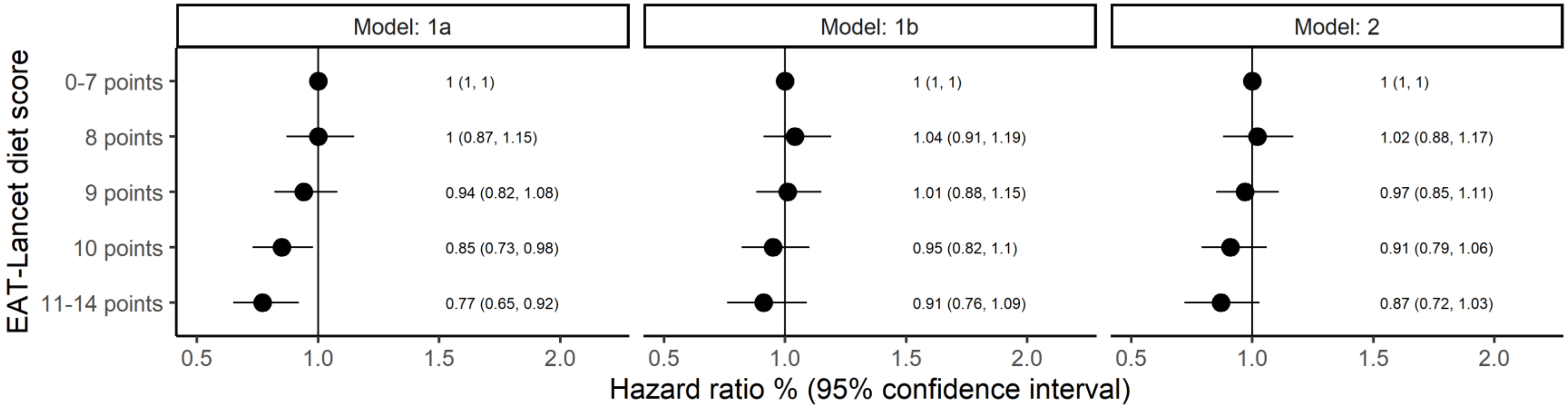


**eFigure 2** Association between adherence to EAT-Lancet diet score and risk of stroke on the relative scale (hazard ratio). Model 1a adjusted for age (underlying time-scale), sex, date of inclusion and age at inclusion (as strata). Model 1b further adjusted for education, smoking status, physical activity, alcohol intake and hormone replacement therapy. Model 2 further adjusted for BMI, waist circumference, history of hypertension, hypercholesterolemia, diabetes and/or acute myocardial infarction. N = 55,016, N cases = 2253.

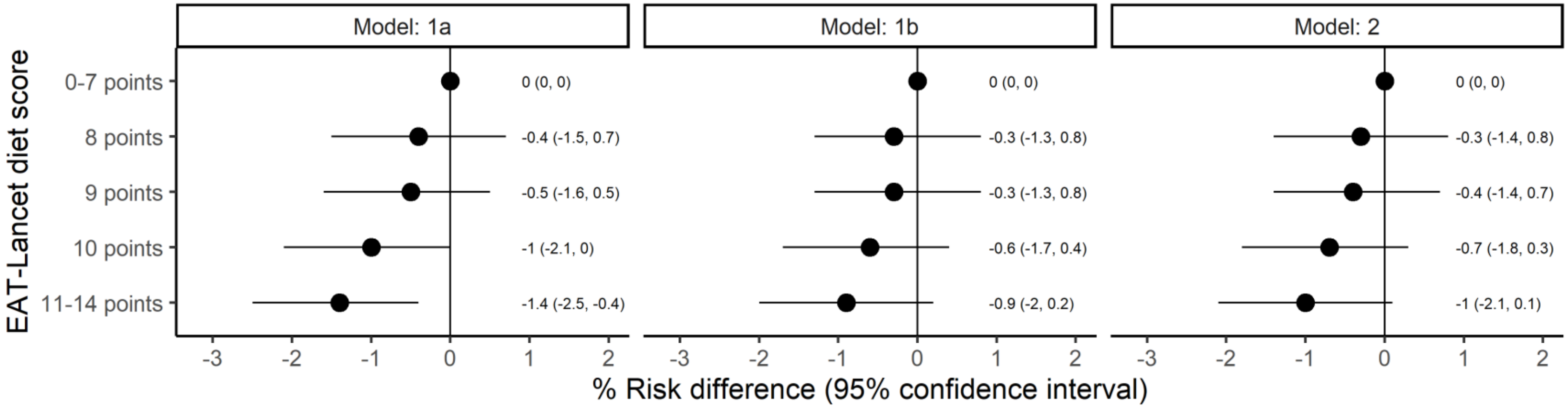


**eFigure 3** Association between adherence to EAT-Lancet diet score and risk of stroke on the absolute scale (risk difference). Model 1a adjusted for age and sex. Model 1b further adjusted for education, smoking status, physical activity, alcohol intake and hormone replacement therapy. Model 2 further adjusted for BMI, waist circumference, history of hypertension, hypercholesterolemia, diabetes and/or acute myocardial infarction. N = 55,016, N cases after 15 years of follow-up = 2251.

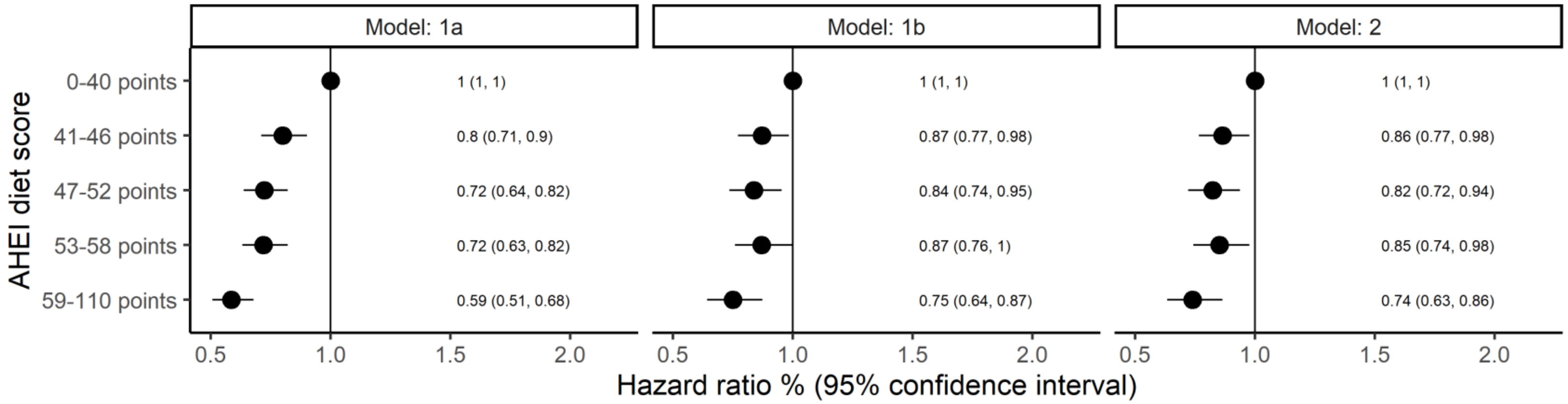


**eFigure 4** Association between adherence to Alternate Healthy Eating Index (AHEI) score and risk of stroke on the relative scale (hazard ratio). Model 1a adjusted for age (underlying time-scale), sex, date of inclusion and age at inclusion (as strata). Model 1b further adjusted for education, smoking status, physical activity, alcohol intake and hormone replacement therapy. Model 2 further adjusted for BMI, waist circumference, history of hypertension, hypercholesterolemia, diabetes and/or acute myocardial infarction. N = 55,016, N cases = 2253.

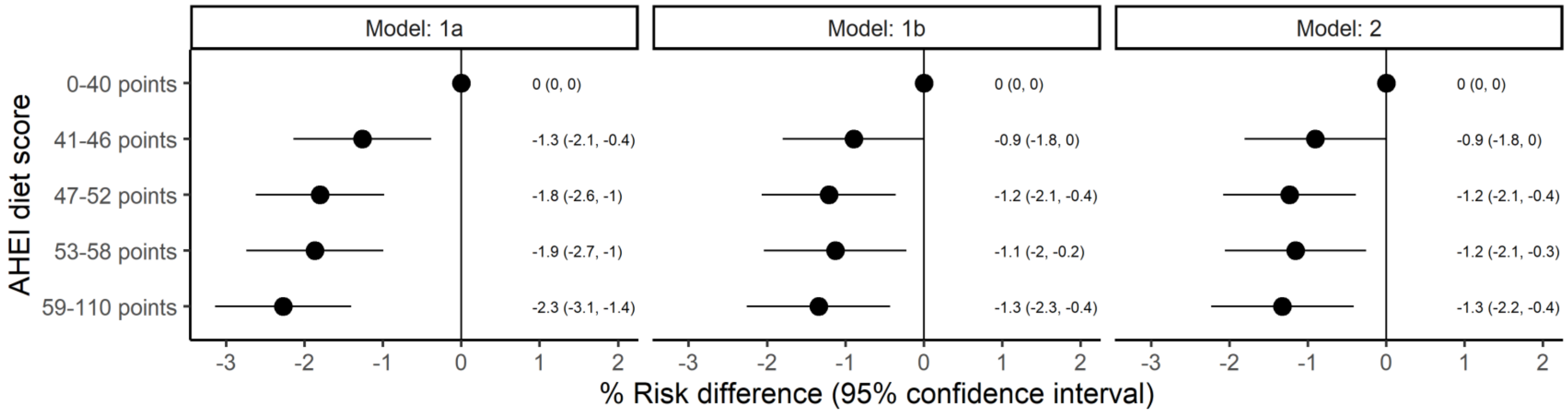


**eFigure 5** Association between adherence to Alternate Healthy Eating Index (AHEI) score and risk of stroke on the absolute scale (risk difference). Model 1a adjusted for age and sex. Model 1b further adjusted for education, smoking status, physical activity, alcohol intake and hormone replacement therapy. Model 2 further adjusted for BMI, waist circumference, history of hypertension, hypercholesterolemia, diabetes and/or acute myocardial infarction. N = 55,016, N cases after 15 years of follow-up = 2251.

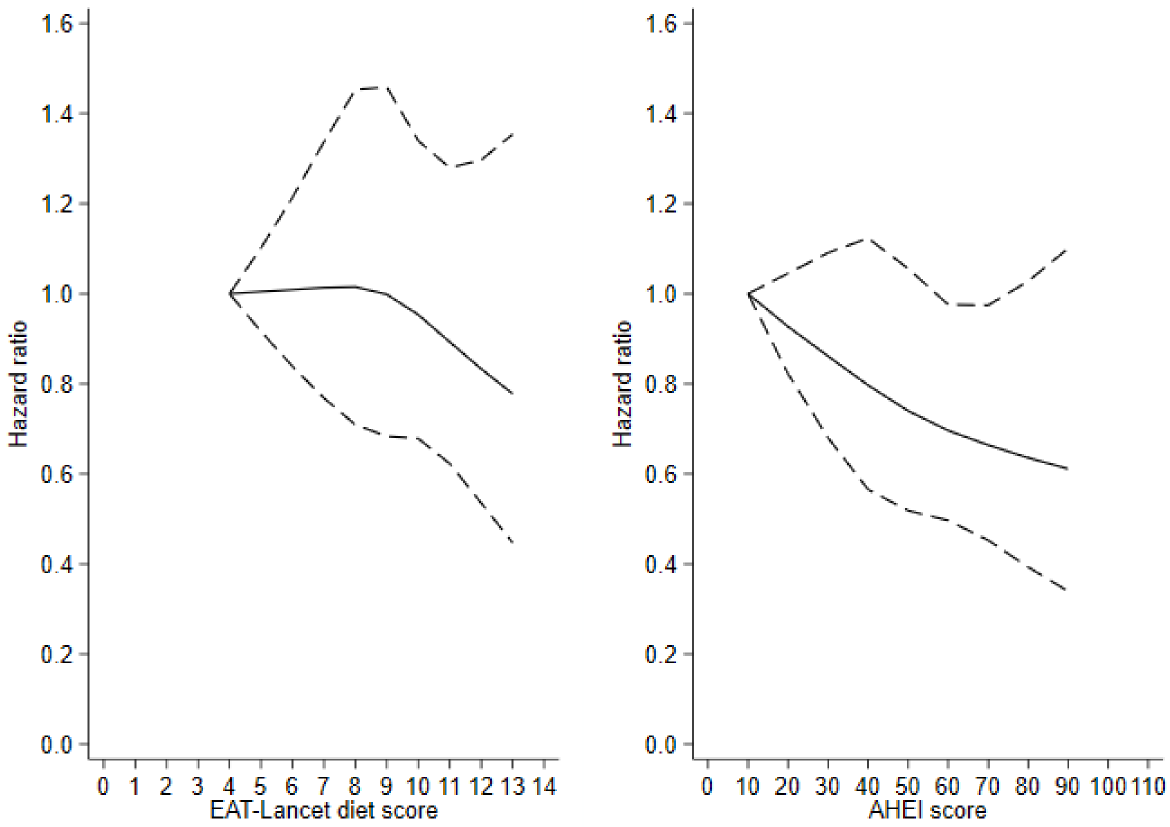


**eFigure 6** Association between adherence to the EAT-Lancet diet score or Alternate Healthy Eating Index (AHEI) score and risk of stroke modelled as restricted cubic splines. Adjusted for age (underlying time-scale), sex, date of inclusion and age at inclusion (as strata), education, smoking status, physical activity, alcohol intake and hormone replacement therapy. N = 55,016, N cases = 2253.

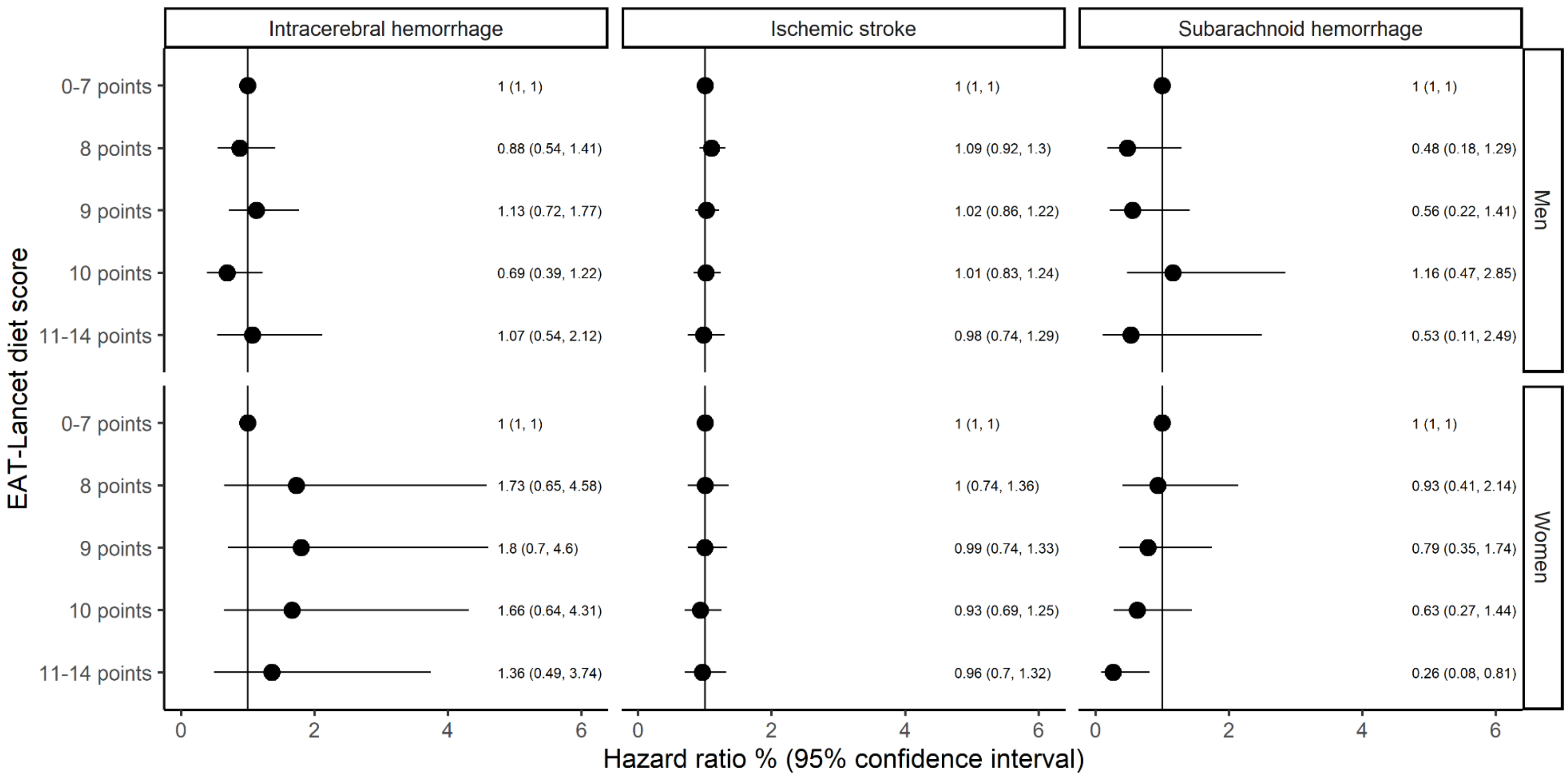


**eFigure 7** Association between adherence to the EAT-Lancet diet score and risk of stroke subtypes in middle-aged men and women. Adjusted for age (underlying time-scale), sex, date of inclusion and age at inclusion (as strata), education, smoking status, physical activity, alcohol intake and hormone replacement therapy. Men: N = 26,208, N intracerebral hemorrhage cases = 160, N ischemic stroke cases = 1148, N subarachnoid hemorrhage cases = 38. Women: N = 28,808, N intracerebral hemorrhage cases = 107, N ischemic stroke cases = 710, N subarachnoid hemorrhage cases = 77.

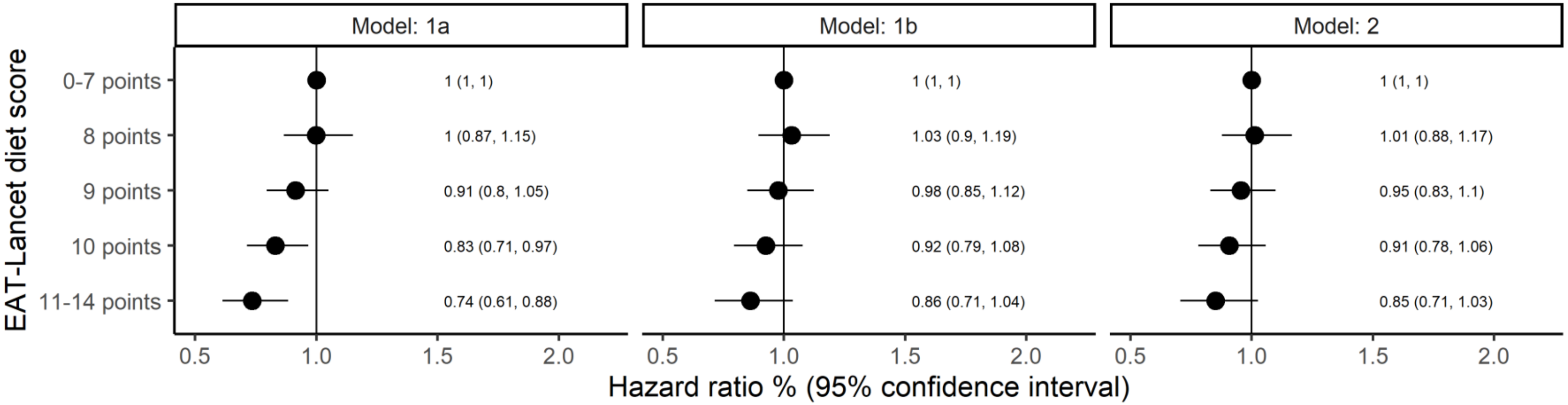


**eFigure 8** Association between adherence to the EAT-Lancet diet score and risk of stroke excluding participants with a history of diabetes or acute myocardial infarction before study start. Model 1a adjusted for age (underlying time-scale), sex, date of inclusion and age at inclusion (as strata). Model 1b further adjusted for education, smoking status, physical activity, alcohol intake and hormone replacement therapy. Model 2 further adjusted for BMI, waist circumference, history of hypertension, hypercholesterolemia, diabetes and/or acute myocardial infarction. N = 53,130, N cases = 2089.
